# Female Infertility Diagnosis and Adult-Onset Psychiatric Conditions: A Matched Cohort Study

**DOI:** 10.1101/2025.06.16.25329704

**Authors:** Khaoula Ben Messaoud, Nina Zaks, Frederick Licciardi, Cecilia Høst Ramlau-Hansen, Linda G. Kahn, Magdalena Janecka

## Abstract

**Study question:** Is there an association between infertility diagnosis and long-term adult-onset psychiatric conditions in women?

**Summary answer:** Infertility diagnosis in women is linked to higher risks of mood disorders, anxiety- and stress-related disorders, and behavioral syndromes with physical components, but not schizophrenia or other psychotic disorders, particularly notable from 9 years after the first infertility diagnosis.

**What is known already:** Infertility, especially in women, is associated with major mental health challenges around the time of diagnosis. However, the long-term connection with a wide range of psychiatric disorders is largely unknown.

**Study design, size, duration:** This study employed a matched-pair design within the UK Biobank (UKB) cohort, including 3,893 females with a diagnosis of infertility and 15,603 matched female controls, totaling 19,496 participants.

**Participants/materials, setting, methods:** Female UKB participants with a diagnosis of infertility were matched to females without the diagnosis in a 1:4 ratio based on year of birth, index of deprivation of their residency area, and primary care data linkage status. The diagnosis of female infertility was identified by the first occurrence of a primary or secondary diagnosis in either primary care or hospital records. Additional analyses explored interactions between infertility diagnosis and both miscarriage and childbearing status on psychiatric conditions.

**Main results and the role of chance:** Diagnosis of infertility was associated with higher risks of mood disorders, anxiety- and stress-related disorders, and behavioral syndromes with physical components, but not with schizophrenia or other psychotic disorders. The most notable increases in the risk of psychiatric diagnoses were observed 9 years after the first infertility diagnosis. No significant interactions were found between infertility diagnosis and either miscarriage or childbearing status on psychiatric conditions. Sensitivity analysis confirmed the robustness of these associations across different data sources for infertility diagnosis and psychiatric condition ascertainment.

**Limitations, reasons for caution:** The study’s limitations include the racial homogeneity and the overall healthier status of the UKB cohort compared to the general UK population, and potential underestimation of associations due to misclassification of subfecund women.

**Wider implications of the findings:** These results emphasize the need for integrated mental health support in infertility care and long-term monitoring of infertility patients for psychiatric risks.

**Study funding/competing interest(s):** The study was conducted using data from the UK Biobank, a prospective cohort study based in the UK. No competing interests were declared.

## Introduction

Infertility is becoming a major health and societal concern, particularly in high-income countries.(Sun *et al*., 2019; Wang *et al*., 2024) Beyond the inability to conceive, infertility frequently imposes an additional burden of morbidity.(Hanson *et al*., 2017; Kong *et al*., 2024; Scime *et al*., 2025) Studies indicate that infertility is associated with significant mental health issues, with women reporting more severe psychological distress, including when compared to men in infertile couples.(Schaller *et al*., 2016; Simionescu *et al*., 2021; Almutawa *et al*., 2023) This psychological toll is further exacerbated by the social stigma of infertility, as well as family-building pressures and expectations.(Kiani *et al*., 2020; Nik Hazlina *et al*., 2022; Bagade *et al*., 2023; Bindeman *et al*., 2025; Peterson *et al*., 2025) Research has consistently shown a higher prevalence of psychiatric disorders such as anxiety and depression among women with infertility, an estimated 36% of whom experience anxiety and 21% of whom experience major depression.(Kiani *et al*., 2020, 2021)

Women’s mental health significantly impacts the outcomes of infertility treatment. Research has shown the time required to achieve success and the likelihood of discontinuing infertility treatment are both closely linked to the mental health status of women undergoing these treatments.(Gameiro *et al*., 2016; Milazzo *et al*., 2016; Purewal *et al*., 2017; Miller *et al*., 2021; Chai *et al*., 2023; Zanettoullis *et al*., 2024) At the same time, the burden and complexity of infertility treatments,(Domar *et al*., 2018) including the effects of hormone therapy, along with the associated direct and indirect financial costs and social pressures, contribute to a higher risk of mental health disorders and more severe symptoms.(Bagade *et al*., 2023) Recognizing the critical role of women’s mental health in infertility care, many specialists are now emphasizing the importance of systematic psychological and/or psychiatric follow-up for women undergoing treatment, with the goal of helping them to manage emotional stress, improve coping strategies, maintain treatment adherence, strengthen relationships, and enhance their overall well-being throughout the IVF process. This approach aims to improve both the outcomes of infertility care and the psychological health of infertile women.(Frederiksen *et al*., 2015; Dube *et al*., 2023)

The nature of the relation between subfecundity (low biological capacity to conceive) and mental health is complex and heterogeneous. Beyond the downstream psychological effects of the inability to conceive and the hormonal unbalance associated with infertility diagnosis and infertility treatments, evidence supports the possibility that female subfecundity and mental health share *upstream* biological causes. Recent findings suggest a genetic correlation between subfecundity and mental health conditions, indicating that certain genetic variants may predispose women to both conditions.(Ma *et al*., 2022; Mao *et al*., 2024; Zeng *et al*., 2024) Other studies suggest the association between infertility and psychiatric outcomes may be moderated by childbearing status(Wdowiak *et al*., 2022; Liu *et al*., 2025) and mediated by cortisol and lipids.(Yang *et al*., 2024)

Previous research has focused on the association between female infertility and mental health conditions primarily during the diagnosis, treatment and immediate postpartum phases.(Egsgaard *et al*., n.d.) However, there is a significant gap in understanding the relation between female infertility and psychiatric conditions over extended timeframes — particularly conditions that manifest prior to, as well as long after, a formal diagnosis of infertility. Such long-term effects could arise due to e.g., the impact of difficulties in conceiving before a formal diagnosis is made, and/or potential factors acting upstream (e.g., genetics) influencing both mental health and female infertility. Furthermore, the evidence linking infertility to various psychiatric disorders remains inconclusive, partly due to the methods used to ascertain psychiatric conditions.(Baldur-Felskov *et al*., 2013)

To address the gaps in understanding the relation between female infertility and psychiatric conditions across the lifespan, this study aims to map the associations between female infertility diagnosis and a wide range of adult-onset psychiatric conditions over the period from 18 years before to 30 years after the diagnosis of infertility. Using a matched-pair design within the UK Biobank (UKB) cohort, we created an ICD-based definition of female infertility and accounted for the potentially differential risks of psychiatric diagnosis in women who, along with an infertility diagnosis, experience miscarriage vs. delivers a live birth. Extensive UKB data allowed us to control for a range of socioeconomic, demographic, and clinical confounders and implement sensitivity analyses to verify the robustness of our results to our analytical decisions.

## Methods

### Data source

The UKB is a longitudinal cohort study based in the UK that included over 500,000 participants aged 40-69 years at recruitment (between 2006 and 2010). The UKB dataset includes information on baseline characteristics (e.g., age at recruitment, month and year of birth, sex, deprivation index at recruitment based on area of residency), lifestyle behaviors, and health-related data collected through questionnaires, interviews, physical measurements, and primary care and hospital records data. Health records linkage is available from before participants’ inclusion in the cohort up to 2023. The cohort profile is detailed elsewhere.(Sudlow *et al*., 2015)

In the current study we used information on the first occurrence of infertility and mental health diagnoses of interest, as well as participant baseline characteristics (demographic and socioeconomic measures). The first occurrence of each diagnosis in UKB participants was ascertained from ICD-9/10 diagnostic data derived from hospital, primary care, and self-reported sources, and assessed using ICD-10 diagnosis codes. Hospitalization data is available for all (100%) and primary care for 45% of UKB participants.

### Study design and infertility diagnosis

We used a matched-pair design within the UKB cohort. The diagnosis of any female infertility was identified by the first occurrence of a primary or secondary diagnosis (ICD-10: N97.0, N97.1, N97.2, N97.3, N97.8, N97.9). Non-diagnosed participants were defined as females without an infertility diagnosis in UKB in both hospital and primary care records. All diagnosed females in the UKB were included in the analyses. Diagnosed and non-diagnosed females were matched in a ratio of 1:4 based on their year of birth, their area Index of Multiple Deprivation (IMD) quintile at enrollment (see the Covariates section), and the presence of primary care linkage. The date of the first infertility diagnosis was used as the index date within each matched group. All women were followed up until their loss to follow-up date in the UKB, which could occur due to reported death, leaving the UK, or withdrawing consent for future linkage in the UKB, or until the end of the study period.

### Psychiatric conditions

Outcomes were first diagnoses of psychiatric conditions (yes/no), categorized according to the ICD-9/10 classification system. Each psychiatric condition was defined by the presence of at least one diagnosis within the ICD-10 code range or their corresponding ICD-9 code (Table 1).

**Table 1:** Psychiatric conditions and related ICD-10 code ranges.

| **Psychiatric conditions group** | **ICD-10** |
| --- | --- |
| Schizophrenia or other psychotic disorders | F20-F29 |
| Mood disorders | F30-F39 |
| Anxiety- and stress-related disorders | F40-F48 |
| Behavioral syndromes with physical components | F50-F59 |

The primary outcomes were the psychiatric conditions listed in Table 1 occurring (a) any time in the study period (18 years before to 30 years after the index date). To increase the temporal resolution of the findings in additional analyses, we also considered psychiatric conditions in the following intervals: (b) before (-18 to 0 years) and after (0 to 30 years) the index date, enabling us to isolate associations that are less likely to reflect the mental health impact of the infertility diagnosis itself; (c) across 6-year intervals relative to the index date, enabling us to track the changes in risk over time. While the analysis of shorter intervals was not possible for all time points due to low frequencies of individuals with both infertility and psychiatric diagnoses, to increase the temporal resolution of the findings where possible, we also computed the estimates for (d) 3- and (e) 1-year intervals.

### Covariates

#### Socio-demographic factors

All socio-demographic variables were ascertained at enrollment in the UKB: year of birth, self-reported race, education, household income, area IMD, and migration status. We adjusted for these variables to account for the temporal changes in the cohort over the enrollment period and social determinants of health influencing the likelihood of receiving both infertility and psychiatric diagnoses.

**Race** was categorized according to the following self-reported categories: White (ref), more than one race, Asian or Asian British including Chinese, and Black or Black British.

**Education** was categorized as: college or university degree; Certificate of Secondary Education (CSE) or equivalent; other professional qualifications; Advanced Level (A/AS) levels or equivalent (ref); Ordinary Level or General Certificate of Secondary Education (O/GCSE) levels or equivalent; National Vocational Qualification (NVQ), Higher National Diploma (HND), or Higher National Certificate (HNC) or equivalent; and "none of the above."

**Household Income** was the family income before tax in British pounds within the following categories: <18,000 (ref); 18,000-30,999; 31,000-51,999; 52,000-100,000; >100,000 and “prefer not to answer”.

The **Index of Multiple Deprivation (IMD)** was calculated by the British government across the local councils across England, Scotland, and Wales. Participants were assigned an IMD score based on their residential address at the time of inclusion. We used the IMD quintile within the regions of England, Scotland, and Wales, using the first quintile (the richest) as the reference.

**Migration status** (yes/no (ref)) was defined by a country of birth different from the UK.

#### Metabolic/endocrine conditions

To account for the associations of metabolic conditions with both infertility and psychiatric conditions, we accounted for the presence of **metabolic/endocrine conditions** prior to the exposure (yes/no (ref)). Presence of a metabolic/endocrine condition was defined based on date of first diagnosis of any ICD-10 code related to obesity (E65-E68), diabetes (E08-E13), thyroid disorders (E01; E02; E04-E07), and other metabolic disorders (E70-E88).

### Reproductive and maternal moderators

The occurrence of **miscarriage** (yes/no (ref)) was defined by the presence of at least one non-elective abortive outcome code (ICD-9: 630-634 and 639; ICD-10: O00-O03, O08) at any time within the study period (-18 to +30 years relative to the index date). This was done to differentiate between cases of infertility where there is evidence of the woman’s ability to establish a pregnancy and cases where there is no such evidence, ensuring the specificity of the female infertility diagnosis and avoiding confusion with the inability to maintain a pregnancy.

**Childbearing status** (yes/no (ref)) was defined as ever having had at least one self-reported live birth. Childbearing is used as a proxy for the severity of infertility (because infertility treatment is covered by national health insurance, we can assume that women with a diagnosis of infertility who did not have a live birth are assumed to have on average, a more severe form of infertility).

### Statistical analysis

#### Main analysis

We estimated associations between female infertility diagnosis and psychiatric outcomes (Table 1) across different time intervals. To account for the matched-pair design, we performed multivariable conditional logistic regressions, yielding odds ratios (ORs) and 95% confidence interval (CIs). In each interval, we included only non-censored females (alive and not lost to follow-up in the UKB) without a prior occurrence of the analyzed psychiatric outcome at the start of the interval. This resulted in a decreasing analytical sample over successive time periods and produced independent estimates across the time periods (i.e., non-cumulative risk estimates). For robust effect estimation, we required that each psychiatric outcome be diagnosed in >5 females with a diagnosis of infertility for analysis across the entire study period and period-specific analyses. All analyses were adjusted for matching variables (primary care data status, IMD, and year of birth), socio-demographic variables (race, education, household income, migration status) and metabolic/endocrine conditions status. Intermediate models were performed to prevent over-adjustment resulting from accounting for metabolic/endocrine conditions. The statistical analysis plan is summarized in Supplementary Table 1. R software (version 4.3.2) was used for all analyses.

#### Secondary analysis

To evaluate the potential moderating roles of lifetime miscarriage and childbearing status, we tested the interaction of associations between those moderators and infertility diagnosis and psychiatric outcomes (mood disorders; anxiety- and stress-related disorders; behavioral syndromes with physical components) at any time, any time before, and any time after, relative to the index date.

#### Sensitivity analysis

Sensitivity analyses were conducted to assess the consistency of our findings across sources of information for (1) the exposure and (2) outcomes by excluding separate information from primary care, hospital, and self-report sources.

## Results

We included 3,893 women with an infertility diagnosis recorded in the UKB as of December 31, 2024, with a mean age at the first diagnosis of 33.4 years. Each woman with an infertility diagnosis was matched to four non-exposed females in the UKB, resulting in an overall sample of 19,496 women. The sample is described in Supplementary Table 2, including the prevalence of each psychiatric condition over the complete study period, the distributions of socio-demographic factors, metabolic conditions, and moderators among exposed and non-exposed females. The number of women in the cohort in each time interval relative to the index date is presented in Supplementary Tables 3-5. The timing of the first diagnosis of psychiatric conditions relative to the index date in the sample is plotted in Supplementary Figure 1.

### Associations between infertility diagnosis and psychiatric conditions

Between the 18 years prior to 30 years following the index date, female infertility diagnosis was associated with an increased risk of mood disorders (OR 1.20, 95% CI, 1.09-1.32); anxiety- and stress-related disorders (OR 1.20, 95% CI, 1.09-1.32); and behavioral syndromes with physical components (OR 1.34, 95% CI, 1.06-1.67), but not with schizophrenia or other psychotic disorders (OR 0.89, 95% CI, 0.47-1.57), adjusting for socio-demographic factors and metabolic and endocrine conditions (Table 2). We observed the same pattern of results when restricting the outcomes to diagnoses of psychiatric conditions occurring after the index date. Infertility diagnosis was not associated with preceding psychiatric diagnoses (Table 3). Intermediate models not adjusted for metabolic/endocrine conditions show similar results (Supplementary Table 6)

**Table 2:** Associations^a^ between Female Infertility Diagnosis and Psychiatric Conditions.

| Psychiatric Conditions | OR<sub>aj</sub> 95% CI |
| --- | --- |
| Schizophrenia, or other psychotic disorders (F20-F29) | 0.89 [0.47-1.57] |
| Mood disorders (F30-F39) | 1.20 [1.09-1.32] |
| Anxiety- and stress-related disorders (F40-F48) | 1.20 [1.09-1.32] |
| Behavioral syndromes with physical components (F50-F59) | 1.34 [1.06-1.67] |

<sup>a</sup> Multivariable conditional logistic regression adjusted for race, education, household income, migration status, metabolic/endocrine conditions status (obesity, diabetes, thyroid disorders and other metabolic disorders), primary care data status, IMD and birth year.

**Table 3:** Associations^a^ between Female Infertility Diagnosis and Psychiatric Conditions by study period window.

| Psychiatric Conditions | OR<sub>aj</sub> 95% CI |  |
| --- | --- | --- |
|  | Any time Before the first infertility diagnosis | Any time After the first infertility diagnosis |
| Schizophrenia, or other psychotic disorders (F20-F29) | 0.59 [0.09-2.17] | 0.97 [0.48-1.81] |
| Mood disorders (F30-F39) | 1.03 [0.86-1.23] | 1.24 [1.11-1.37] |
| Anxiety- and stress-related disorders (F40-F48) | 1.09 [0.85-1.37] | 1.20 [1.09-1.33] |
| Behavioral syndromes with physical components (F50-F59) | 1.07 [0.73-1.53] | 1.56 [1.17-2.06] |

<sup>a</sup> Multivariable conditional logistic regression adjusted for race, education, household income, migration status, metabolic/endocrine conditions status (obesity, diabetes, thyroid disorders and other metabolic disorders), primary care data status, IMD and birth year.

The associations between female infertility diagnosis and each of the psychiatric outcome’s conditions over time (every 6 years before/after the index date) are presented in Figure 1 and Supplementary Table 6. Female infertility diagnosis was associated with mood disorders and anxiety- and stress-related disorders beginning 12 years after the infertility diagnosis (at 12, 18, 24, and 30 years); and with behavioral syndromes with physical components beginning 18 years after infertility diagnosis (at 18, 24, and 30 years; Supplementary Table 6). We did not observe higher risk of psychiatric diagnosis within the first six years of infertility diagnosis. Associations in 3- and 1-year intervals are presented in Supplementary Tables 7-8 and Supplementary Figures 2-3. The analyses conducted at three-year intervals revealed that the associations became significant as early as nine years after the index date.

**Figure 1:**
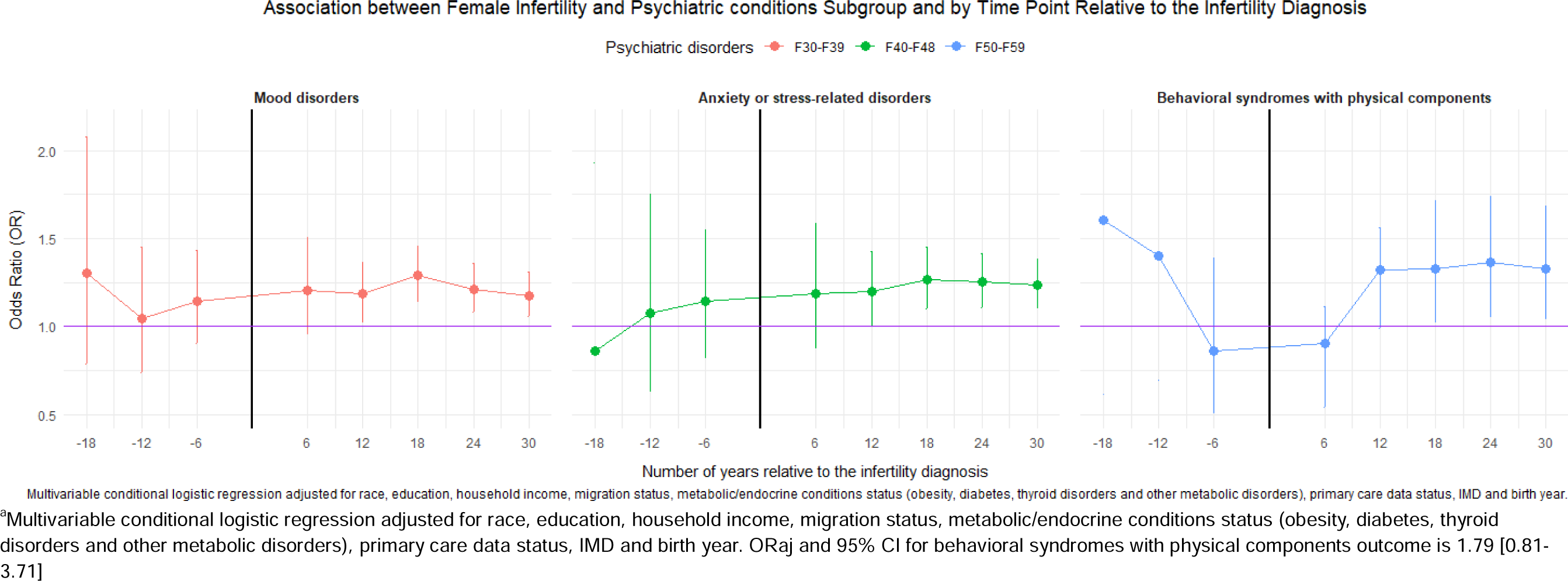
Associations^a^ between female infertility diagnosis and psychiatric conditions in 6-year intervals relative to the infertility diagnosis.

### Interactions with miscarriage and childbearing

There were no interactions between female infertility diagnosis and either miscarriage or childbearing status on psychiatric conditions any time, before, or after the first diagnosis of infertility (Table 4).

**Table 4:**
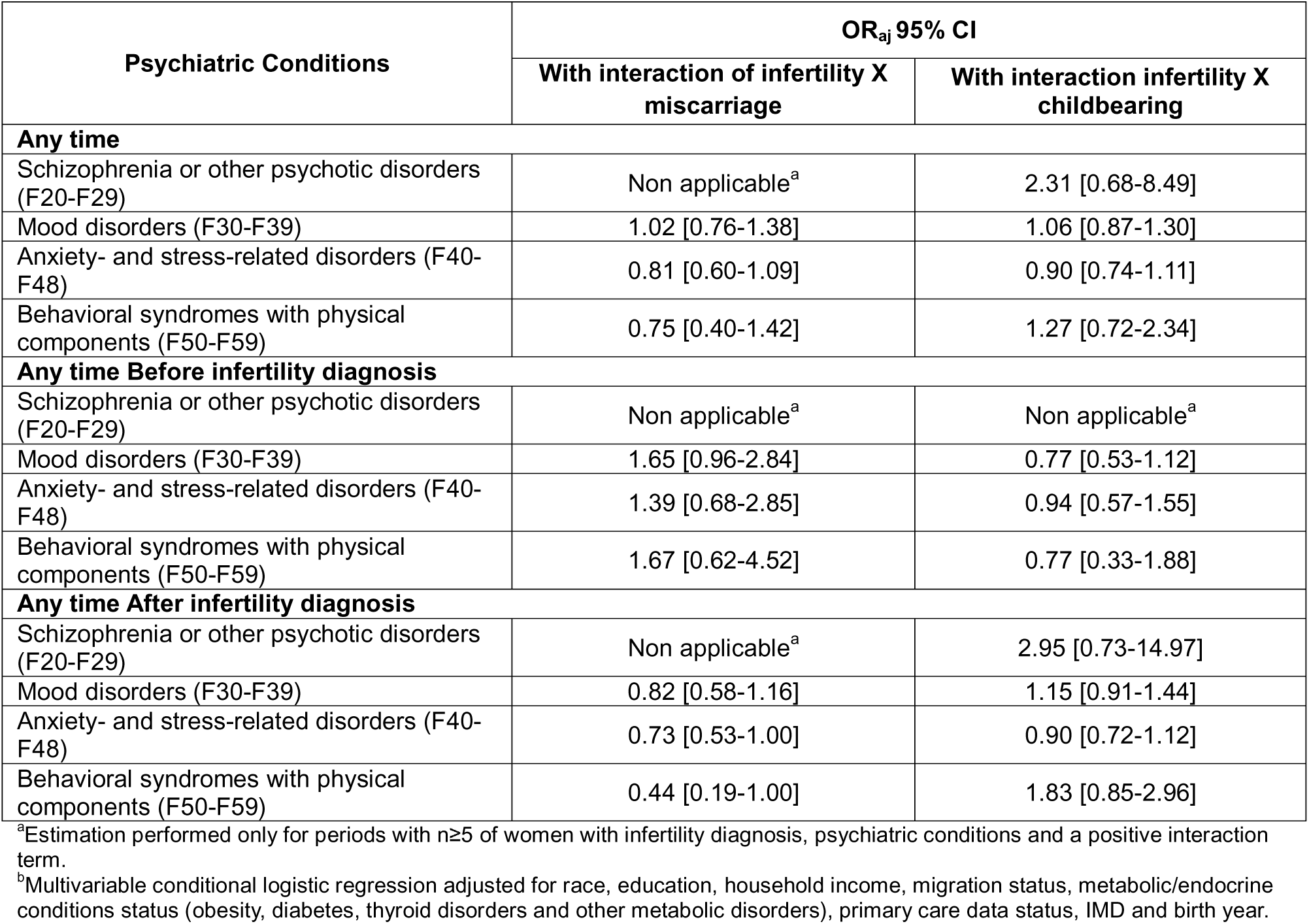
Associations^a,b^ between Female Infertility Diagnosis, Miscarriage, Childbearing, and Psychiatric Conditions.

### Sensitivity analysis

The sensitivity analysis testing whether the associations between female infertility diagnosis and psychiatric conditions change depending on the source of information (hospitalization, primary care, self-report), showed similar findings compared with the main analysis for both the outcomes in the entire study period and by time point (Supplementary Table 9).

## Discussion

Our study shows that infertility diagnosis is associated with multiple, long-term mental health challenges. Specifically, female infertility diagnosis is significantly associated with higher risks of mood disorders, anxiety- and stress-related disorders, and behavioral syndromes with physical components starting 9 years following the infertility diagnosis up to at least 30 years post-diagnosis. There was no evidence that these associations were present in the period preceding or in the first several years following infertility diagnosis, or that they were moderated by the experience of miscarriage or childbearing. Our results were not affected by any over-adjustment related to metabolic variables and were robust to potential bias involving the source of diagnostic data.

Our findings indicate higher risk of adult-onset psychiatric conditions in women diagnosed with infertility, except for schizophrenia or other psychotic disorders. These results are consistent with previous research that has demonstrated an association between female infertility and long-term risk of mood disorders, as well as a genetic correlation between infertility and depression.(Vikström *et al*., 2015; Hanson *et al*., 2017; Ma *et al*., 2022; Mao *et al*., 2024; Zeng *et al*., 2024) Furthermore, we identified a link between female infertility diagnosis and long-term risk of anxiety and stress-related disorders. This association has been investigated in only one previous study based in Denmark, which found no link between female fertility problems and hospitalization for anxiety. The disparate results between our study and the Danish study could have arisen due to differences in the ascertainment of female infertility (ICD diagnosis of infertility vs. not giving birth after infertility evaluation) and psychiatric conditions (anxiety ascertained through both primary care and hospitalization data, vs. just hospitalization).(Baldur-Felskov *et al*., 2013) Our findings are consistent with the other studies reporting an association between reproductive disorders linked with subfecundity, such as polycystic ovary syndrome, and the risk of anxiety.(Zaks *et al*., 2023; Infante-Cano *et al*., 2024) We did not find an association between female infertility diagnosis and schizophrenia or other psychotic disorders (0.89 [0.47-1.57]), possibly due to the limited sample size, as only 13 women with infertility diagnosis in the UKB developed these conditions during the study period. In contrast, the Danish study reported a link between not giving birth after infertility evaluation and hospitalization for schizophrenia and psychoses.(Baldur-Felskov *et al*., 2013; Hanson *et al*., 2017)

Behavioral syndromes with physical components were the psychiatric conditions most strongly associated with female infertility diagnosis in our study, with a lifetime odds ratio of 1.34 [1.06-1.67]. Behavioral syndromes with physical components include diagnoses related to physiological disturbances, such as eating or sleep disorders and the abuse of non-dependence-producing substances. These conditions may involve metabolic and hormonal imbalances, which are also linked to infertility. For example, a systematic review found that sleep disorders are associated with female infertility, although the studies reviewed investigated primarily sleep disorders during treatment or while attempting to conceive.(Li *et al*., 2024) Additionally, a sibling comparison study using the Swedish register showed a link between polycystic ovarian syndrome (PCOS) diagnosis – often associated with infertility - and the risk of sleeping, sexual, and eating disorders, even after accounting for family and early-life factors. The authors suggest that factors shared by the siblings, such as shared environmental influences, lifestyle choices, or genetic factors, could affect both PCOS and these related conditions.(Boldis *et al*., 2024)

Female infertility diagnosis was associated with non-psychotic psychiatric conditions starting 9 years post-diagnosis. In our sample, the mean age of women at their first infertility diagnosis was 33.4 years, indicating that the association was significant in women past, on average, 42-45 years of age. The relatively late onset of psychiatric diagnoses may indicate that such effects increase around the transition to menopause, when women experience significant hormonal changes. Alternatively, this could also be attributed to delays in diagnosing psychiatric conditions. Delays in diagnosis may also explain our finding of an absence of association between psychiatric conditions prior to infertility diagnosis, which contrasts with previous studies’ findings of a genetic correlation between female infertility and depression,(Ma *et al*., 2022; Mao *et al*., 2024; Zeng *et al*., 2024) anxiety spectrum, and stress-related disorders.(Ma *et al*., 2022) This inconsistency may also be due to a lack of statistical power, as our sample includes a relatively small number of women with these outcomes occurring during the period before and the first years after the infertility diagnosis, which can explain the larger confidence intervals.

Interactions between female infertility diagnosis and miscarriage or childbearing on the adult-onset psychiatric conditions were all non-significant, suggesting that these reproductive and maternal factors—specifically, the severity of infertility as indicated by childbearing and the inability to maintain a pregnancy as indicated by miscarriage—do not moderate associations of infertility diagnosis with the development of psychiatric conditions. However, as the confidence intervals around many estimates were wide, this should be tested in larger datasets.

Analyzing associations between infertility diagnosis and broad categories of adult-onset psychiatric conditions across the reproductive life span enabled us to expand upon the existing literature by considering lags and not presuming a temporal precedence of the infertility diagnosis. The robustness of our findings is supported by the high quality of the UKB dataset, which includes extensive and detailed population data, medical history, as well as prospective follow-up. Additionally, the methods and study design we employed help minimize selection bias and allow for adjustments across a range of socio-economic and metabolic variables. Our findings were consistent with analyses conducted globally and were shown to be robust to the source of medical information. Despite our rigorous study design and comprehensive follow-up data of the UKB, our study has some limitations. First, our cohort was relatively homogeneous in terms of race, as 88% of the sample analyzed self-reported as White. Additionally, the UKB cohort is overall healthier than the general population in the UK(Sudlow *et al*., 2015). Furthermore, associations in our analysis may be underestimated, as the use of female infertility diagnosis can misclassify women with subfecundity who did not try to get pregnant, did not met the full clinical criteria, or did not receive a diagnosis due to e.g., lower access to healthcare.

## Conclusion

Our research shows that female infertility diagnosis is associated with a higher risk of psychiatric conditions. This underscores the critical need for integrating mental health support into infertility care and for the long-term monitoring of psychiatric risks among women who receive an infertility diagnosis, highlighting significant implications for both clinical practice and public health policy. Ensuring that women diagnosed with infertility receive early and comprehensive mental health care is crucial. Public health initiatives should aim to raise awareness about the psychological burden of infertility diagnosis and promote early intervention strategies to mitigate long-term psychiatric risks. Further research is warranted to explore potential underlying biological mechanisms shared by female subfecundity and psychiatric morbidity to develop targeted interventions.

## Author’s roles

Dr. Ben Messaoud had full access to all the data in the study and takes responsibility for the integrity of the data and the accuracy of the data analysis.

*Concept and design*: Ben Messaoud and Janecka

*Acquisition, analysis, or interpretation of data*: Ben Messaoud and Janecka

*Drafting of the manuscript*: Ben Messaoud and Janecka.

*Statistical analysis:* Ben Messaoud.

*Critical review of the manuscript for important intellectual content:* Ben Messaoud, Zaks, Licciardi, Ramlau-Hansen, Kahn, Janecka

*Administrative, technical, or material support:* Ben Messaoud, Janecka and Zaks.

*Supervision:* Janecka

## Data Access, Responsibility, and Analysis

UK Biobank received ethical approval from the Research Ethics Committee (REC reference for UK Biobank is 11/NW/0382). Informed consent was obtained from all subjects involved in the study.

## Data Sharing Statement

UKB data are available for bona fide researchers (https://www.ukbiobank.ac.uk).

