## Supplemental files for "Female Infertility Diagnosis and Adult-Onset Psychiatric Conditions: A Matched Cohort Study"

**Supplementary Table 1: Summary of the statistical analysis.**

| **Psychiatric Conditions** | **Anytime/ Any time before/ Any time after** | **Time points by 6 years**  (-18, -12, -6, 6, 12, 12, 18, 24, 30) | **Time points by 3 years**  (-18, -15, -12, -9, -6, -3, 3, 6, 9, 12, 15, 18, 24, 27, 30) | **Time points by year**  (-3, -2, -1, 1, 2, 3) |
| --- | --- | --- | --- | --- |
| **Main analysis** | | | | |
| Schizophrenia, and other psychotic disorders | Model (1) | **X** | **X** | **X** |
| Mood disorders | Model (2) | Model (2) | Model (2) | Model (2) |
| Anxiety- or stress-related disorders | Model (3) | Model (3) | Model (3) | Model (3) |
| Behavioral syndromes with physical components | Model (4) | Model (4) | Model (4)^a^ | Model (4)^a^ |
| **Secondary analysis** | | | | |
| Schizophrenia, and other psychotic disorders | Model interaction infertility X Miscarriage (1a)  Model interaction infertility X childbearing (1b) | **X** | **X** | **X** |
| Mood disorders | Model interaction infertility X Miscarriage (2a)  Model interaction infertility X childbearing (2b) | **X** | **X** | **X** |
| Anxiety- or stress-related disorders | Model interaction infertility X Miscarriage (3a)  Model interaction infertility X childbearing (3b) | **X** | **X** | **X** |
| Behavioral syndromes with physical components | Model interaction infertility X Miscarriage (4a)  Model interaction infertility X childbearing (4b) | **X** | **X** | **X** |
| **Sensitivity analysis for the exposition^2^** | | | | |
| Mood disorder | | | | |
| Self-report | Model (2) | Model (2) | **X** | **X** |
| Hospital | Model (3) | Model (3) | **X** | **X** |
| Primary care | Model (4) | Model (4) | **X** | **X** |
| Anxiety- or stress-related disorders | | | | |
| Self-report | Model (2) | Model (2) | **X** | **X** |
| Hospital | Model (3) | Model (3) | **X** | **X** |
| Primary care | Model (4) | Model (4) | **X** | **X** |
| Mood disorder | | | | |
| Self-report | Model (2) | Model (2) | **X** | **X** |
| Hospital | Model (3) | Model (3) | **X** | **X** |
| Primary care | Model (4) | Model (4) | **X** | **X** |

^a^ Estimation performed only for periods with n≥5 of women with infertility diagnosis, psychiatric conditions.

**Supplementary Table 2: Covariates and psychiatric conditions according to infertility diagnosis status.**

| **Variables** | **With infertility diagnosis** | | **Without infertility diagnosis** | |
| --- | --- | --- | --- | --- |
|  | **N=****3893** | **%** | **N=15572** | **%** |
| **I****ndex of Multiple Deprivation quintile^a^** | | | | |
| 1^st^ quintile (richest) | 808 | 20.76 | 3232 | 20.76 |
| 2^nd^ quintile | 804 | 20.65 | 3216 | 20.65 |
| 3^rd^ quintile | 829 | 21.29 | 3316 | 21.29 |
| 4^th^ quintile | 795 | 20.42 | 3180 | 20.42 |
| 5^th^ quintile (poorest) | 657 | 16.88 | 2628 | 16.88 |
| **Income** | | | | |
| Less than 18,000 | 499 | 12.82 | 2481 | 15.93 |
| 18,000 to 30,999 | 678 | 17.42 | 3159 | 20.29 |
| 31,000 to 51,999 | 984 | 25.28 | 3745 | 24.05 |
| 52,000 to 100,000 | 944 | 24.25 | 3189 | 20.48 |
| Greater than 100,000 | 261 | 6.70 | 823 | 5.29 |
| Prefer not to answer | 383 | 9.84 | 1443 | 9.27 |
| Missing value | 144 | 3.70 | 732 | 4.70 |
| **Degree** | | | | |
| A levels/AS levels or equivalent | 98 | 2.52 | 489 | 3.14 |
| College or University degree | 521 | 13.38 | 1733 | 11.13 |
| CSEs or equivalent | 376 | 9.66 | 1555 | 9.99 |
| None of the above | 336 | 8.63 | 1623 | 10.42 |
| NVQ or HND or HNC or equivalent | 534 | 13.72 | 2229 | 14.31 |
| O levels/GCSEs or equivalent | 719 | 18.47 | 3044 | 19.55 |
| Other professional qualifications (nursing, teaching) | 1258 | 32.31 | 4697 | 30.16 |
| Missing value | 51 | 1.31 | 202 | 1.30 |
| **Race** | | | | |
| White | 3447 | 88.54 | 13968 | 89.70 |
| Asian or Asian British | 179 | 4.60 | 738 | 4.74 |
| Black or Black British | 31 | 0.80 | 95 | 0.61 |
| More than one race | 168 | 4.32 | 536 | 3.44 |
| Other ethnic groups | 49 | 1.26 | 156 | 1.00 |
| Missing value | 19 | 0.49 | 79 | 0.51 |
| **Migration status** | | | | |
| No | 3527 | 90.60 | 14205 | 91.22 |
| Yes | 358 | 9.20 | 1339 | 8.60 |
| Missing value | 8 | 0.21 | 28 | 0.18 |
| **Metabolic/endocrine conditions** | | | | |
| No | 2741 | 70.41 | 11149 | 71.60 |
| Yes | 1152 | 29.59 | 4423 | 28.40 |
| **Childbearing status any time^b^** |  | | | |
| No | 1454 | 37.35 | 3486 | 22.39 |
| Yes | 2428 | 62.37 | 12047 | 77.36 |
| Missing value | 11 | 0.28 | 39 | 0.25 |
| **Miscarriage any time^b^** | | | | |
| No | 3310 | 85.02 | 14692 | 94.35 |
| Yes | 583 | 14.95 | 880 | 5.65 |
| **Schizophrenia and other psychotic disorders (F20-F29) any time^b^** | | | | |
| No | 3880 | 99.67 | 15498 | 99.52 |
| Yes | 13 | 0.33 | 74 | 0.48 |
| **Mood disorders (F30-F39) any time^b^** | | | | |
| No | 3128 | 80.35 | 12797 | 82.18 |
| Yes | 765 | 19.65 | 2775 | 17.82 |
| **Anxiety- or stress-related disorders (F40-F48) any time^b^** | | | | |
| No | 3133 | 80.48 | 12868 | 82.64 |
| Yes | 760 | 19.52 | 2704 | 17.36 |
| **Behavioral syndromes with physical components (F50-F59) any time^b^** | | | | |
| No | 3785 | 97.23 | 15240 | 97.87 |
| Yes | 108 | 2.77 | 332 | 2.13 |
| **Primary care linkage^a^** | | | | |
| No | 1020 | 26.20 | 4080 | 26.20 |
| Yes | 2873 | 73.80 | 11492 | 73.80 |
| **Geographical area** | | | | |
| England | 3262 | 83.79 | 13177 | 84.62 |
| Scotland | 433 | 11.12 | 1443 | 9.27 |
| Wales | 198 | 5.09 | 952 | 6.11 |
| **Numeric variables mean (±SD, median; Q1-Q3)** | | | | |
| Age at recruitment | 51.0(±7.8, 49.0; 44.0-57.0) | | 51.1(±7.8, 49.0; 44.0-57.0) | |
| Year of birth^a^ | 1957.1(±7.8, 1959; 1951-1964) | | 1957.1(±7.8, 1959; 1951-1964) | |
| Age at the first diagnosis of infertility | 33.4(±6.2, 33.6; 28.8-37.9) | | Non applicable | |
| Age at first diagnosis of schizophrenia and other psychotic disorders | 49.2(±14.9, 50.8; 41.5-58.3) | | 49.2(±14.9, 50.8; 36.0-59.4) | |
| Age at first diagnosis of mood disorders | 44.0(±13.0, 44.5; 35.2-52.9) | | 44.2(±13.5, 44.5; 35.6-53.1) | |
| Age at first diagnosis of anxiety- or stress-related disorders | 48.0(±12.4, 48.3; 40.7-55.8) | | 48.8(±12.8, 49.0; 41.0-56.7) | |
| Age at first diagnosis of behavioral syndromes with physical components | 37.1(±13.0, 36.8; 29.1-45.3) | | 36.3(±12.6, 34.6; 27.7-44.4) | |

^a^Matching variables

^b^Unrestricted follow-up, encompassing the entire duration of follow-up available in the UK Biobank

**Supplementary Table 3: Number of women in the cohort with both infertility diagnosis and psychiatric conditions by time intervals (6-year intervals) relative to infertility diagnosis.**

| **Time Point (years)** | **-18** | **-12** | **-6** | **6** | **12** | **18** | **24** | **30** |
| --- | --- | --- | --- | --- | --- | --- | --- | --- |
| Schizophrenia and other psychotic disorders (F20-F29) | 0 | 1 | 1 | 1 | 2 | 2 | 3 | 1 |
| Mood disorders (F30-F39) | 8 | 23 | 55 | 192 | 136 | 102 | 79 | 67 |
| Anxiety- or stress-related disorders (F40-F48) | 2 | 13 | 34 | 138 | 136 | 131 | 104 | 84 |
| Behavioral syndromes with physical components (F50-F59) | 3 | 5 | 7 | 48 | 13 | 11 | 5 | 9 |

**Supplementary Table 4: Number of women in the cohort with both infertility diagnosis and psychiatric conditions by time intervals (3-year intervals) relative to infertility diagnosis.**

| **Time Point (years)** | **-18** | **-15** | **-12** | **-9** | **-6** | **-3** | **3** | **6** | **9** | **12** | **15** | **18** | **21** | **24** | **27** | **30** |
| --- | --- | --- | --- | --- | --- | --- | --- | --- | --- | --- | --- | --- | --- | --- | --- | --- |
| Schizophrenia and other psychotic disorders (F20-F29) | 0 | 1 | 0 | 0 | 1 | 0 | 0 | 1 | 1 | 1 | 0 | 2 | 2 | 1 | 0 | 1 |
| Mood disorders (F30-F39) | 8 | 10 | 13 | 23 | 32 | 28 | 100 | 64 | 71 | 65 | 54 | 48 | 43 | 36 | 42 | 25 |
| Anxiety- or stress-related disorders (F40-F48) | 2 | 5 | 8 | 14 | 20 | 18 | 67 | 53 | 64 | 72 | 65 | 66 | 49 | 55 | 53 | 31 |
| Behavioral syndromes with physical components (F50-F59) | 3 | 1 | 4 | 7 | 0 | 10 | 25 | 13 | 9 | 4 | 4 | 7 | 2 | 3 | 4 | 5 |

**Supplementary Table 5: Number** **of women in the cohort with both infertility diagnosis and psychiatric conditions by time intervals (1-year intervals) relative to infertility diagnosis.**

| **Time Point (years)** | **-3** | **-2** | **-1** | 1 | 2 | 3 |
| --- | --- | --- | --- | --- | --- | --- |
| Schizophrenia and other psychotic disorders (F20-F29) | 0 | 0 | 0 | 0 | 0 | 0 |
| Mood disorders (F30-F39) | 28 | 10 | 12 | 30 | 22 | 26 |
| Anxiety- or stress-related disorders (F40-F48) | 18 | 8 | 11 | 23 | 12 | 13 |
| Behavioral syndromes with physical components (F50-F59) | 10 | 3 | 1 | 6 | 11 | 4 |

**Supplementary Figure 1: Timing of psychiatric conditions diagnosis relative to infertility diagnosis.**

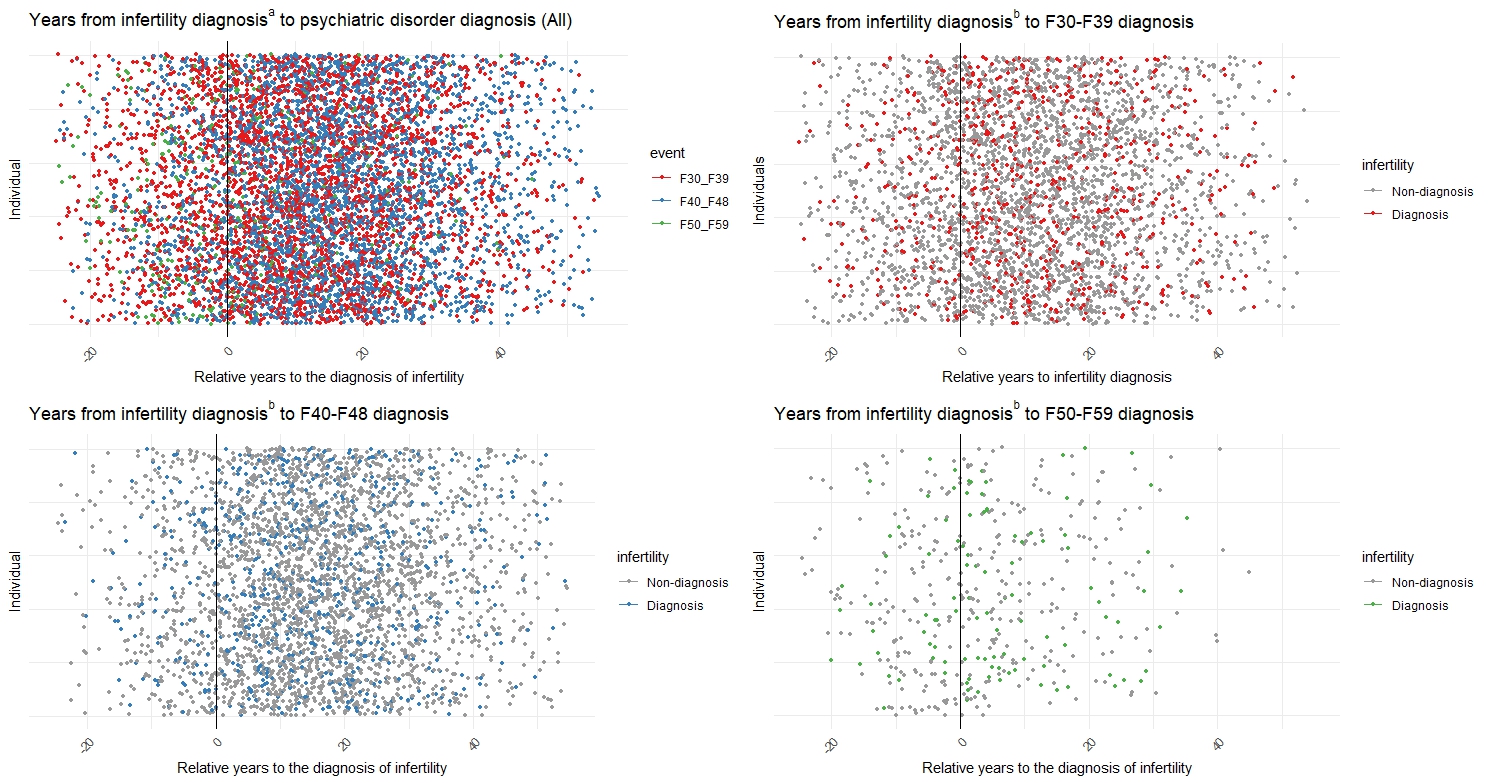

^a^ Women with infertility diagnosis are represented

^b^ The plot includes both women with and without infertility diagnosis. For women without infertility diagnosis, the date of the first infertility diagnosis within the matching subgroup was used for representation.

**Supplementary Table 6: Associations^a,b^ between Female Infertility Diagnosis and Psychiatric Conditions by study period window.**

| **Psychiatric Conditions** | **OR_aj_ 95% CI** | |
| --- | --- | --- |
|  | **Intermediate Model 1^a^** | **Intermediate Model 2^b^** |
| **Any time** | | |
| Schizophrenia and other psychotic disorders (F20-F29) | 0.70 [0.37-1.22] | 0.90 [0.47-1.58] |
| Mood disorders (F30-F39) | 1.13 [1.03-1.23] | 1.21 [1.10-1.33] |
| Anxiety- or stress-related disorders (F40-F48) | 1.16 [1.06-1.27] | 1.21 [1.10-1.33] |
| Behavioral syndromes with physical components (F50-F59) | 1.31 [1.05-1.63] | 1.34 [1.07-1.68] |
| **Any time Before infertility diagnosis** | | |
| Schizophrenia and other psychotic disorders (F20-F29) | 0.50[0.08-1.76] | 0.61 [0.09-2.23] |
| Mood disorders (F30-F39) | 1.00 [0.84-1.19] | 1.04 [0.87-1.25] |
| Anxiety- or stress-related disorders (F40-F48) | 1.07 [0.85-1.34] | 1.09 [0.86-1.38] |
| Behavioral syndromes with physical components (F50-F59) | 1.04 [0.71-1.48] | 1.07 [0.73-1.53] |
| **Any time After infertility diagnosis** | | |
| Schizophrenia and other psychotic disorders (F20-F29) | 0.76 [0.38-1.39] | 0.98 [0.48-1.82] |
| Mood disorders (F30-F39) | 1.16 [1.05-1.28] | 1.25 [1.12-1.38] |
| Anxiety- or stress-related disorders (F40-F48) | 1.15 [1.04-1.27] | 1.21 [1.10-1.34] |
| Behavioral syndromes with physical components (F50-F59) | 1.53 [1.16-2.01] | 1.57 [1.17-2.07] |

^a^ Multivariable conditional logistic regression adjusted for matching variables (primary care data status, IMD and birth year).

^b^ Multivariable conditional logistic regression adjusted for matching variables (primary care data status, IMD and birth year), and sociodemographic variables (race, education, household income, migration status).

**Supplementary Table 7: Association^a,b^ between infertility diagnosis and psychiatric conditions by subgroup and by time point (6-year intervals) relative to the infertility diagnosis.**

|  | **Mood disorders F30-F39** | | **Anxiety- or stress-related disorders F40-F48** | | **Behavioral syndromes with physical components F50-F59** | |
| --- | --- | --- | --- | --- | --- | --- |
| **Time point** | **OR_adj_** | **95% CI** | **OR_adj_** | **95% CI** | **OR_adj_** | **95% CI** |
| **-18** | 1.30 | [0.79-2.08] | 0.86 | [0.32-1.93] | 1.61 | [0.62-3.75] |
| **-12** | 1.04 | [0.74-1.45] | 1.08 | [0.63-1.75] | 1.40 | [0.69-2.66] |
| **-6** | 1.14 | [0.90-1.43] | 1.14 | [0.83-1.55] | 0.86 | [0.51-1.39] |
| **6** | 1.21 | [0.96-1.50] | 1.19 | [0.88-1.59] | 0.90 | [0.54-1.11] |
| **12** | 1.19 | [1.03-1.37] | 1.20 | [1.00-1.43] | 1.32 | [0.99-1.56] |
| **18** | 1.29 | [1.14-1.46] | 1.27 | [1.10-1.45] | 1.33 | [1.02-1.72] |
| **24** | 1.21 | [1.08-1.36] | 1.25 | [1.11-1.41] | 1.37 | [1.06-1.74] |
| **30** | 1.18 | [1.05-1.31] | 1.23 | [1.10-1.38] | 1.33 | [1.04-1.68] |

^a^ Multivariable conditional logistic regression adjusted for race, education, household income, migration status, metabolic/endocrine conditions status (obesity, diabetes, thyroid disorders and other metabolic disorders), primary care data status, IMD and birth year.

^b^ Estimation performed only for time intervales with n≥5 of women with both infertility diagnosis and psychiatric conditions.

**Supplementary Table 8: Association^a,b^ between female infertility and psychiatric conditions by subgroup and by time point (3-year intervals) relative to the infertility diagnosis.**

|  | **Mood disorders F30-F39** | | **Anxiety- or stress-related disorders F40-F48** | | **Behavioral syndromes with physical components F50-F59** | |
| --- | --- | --- | --- | --- | --- | --- |
| **Time point** | **OR_adj_** | **95% CI** | **OR_adj_** | **95% CI** | **OR_adj_** | **95% CI** |
| **-18** | 1.3 | [0.79-2.08] | 0.86 | [0.32-1.93] | Non applicable^b^ | Non applicable^b^ |
| **-15** | 1.00 | [0.65-1.47] | 1.02 | [0.5-1.91] | Non applicable^b^ | Non applicable^b^ |
| **-12** | 1.04 | [0.74-1.45] | 1.08 | [0.63-1.75] | Non applicable^b^ | Non applicable^b^ |
| **-9** | 1.06 | [0.8-1.38] | 1.14 | [0.76-1.67] | 1.25 | [0.72-2.05] |
| **-6** | 1.14 | [0.9-1.43] | 1.14 | [0.83-1.55] | Non applicable^b^ | Non applicable^b^ |
| **-3** | 1.06 | [0.86-1.29] | 1.11 | [0.84-1.45] | 1.02 | [0.66-1.52] |
| **3** | 1.13 | [0.92-1.38] | 1.14 | [0.86-1.48] | 1.13 | [0.75-1.66] |
| **6** | 1.16 | [0.99-1.36] | 1.17 | [0.95-1.42] | 1.21 | [0.88-1.65] |
| **9** | 1.23 | [1.07-1.41] | 1.19 | [0.99-1.41] | 1.32 | [1.05-1.74] |
| **12** | 1.25 | [1.1-1.42] | 1.26 | [1.08-1.46] | Non applicable^b^ | Non applicable^b^ |
| **15** | 1.28 | [1.13-1.44] | 1.27 | [1.11-1.46] | Non applicable^b^ | Non applicable^b^ |
| **18** | 1.27 | [1.13-1.42] | 1.24 | [1.09-1.41] | 1.33 | [1.15-71] |
| **21** | 1.22 | [1.09-1.36] | 1.26 | [1.12-1.42] | Non applicable^b^ | Non applicable^b^ |
| **24** | 1.20 | [1.07-1.33] | 1.2 | [1.07-1.35] | Non applicable^b^ | Non applicable^b^ |
| **27** | 1.18 | [1.06-1.32] | 1.24 | [1.11-1.38] | Non applicable^b^ | Non applicable^b^ |
| **30** | 1.18 | [1.07-1.31] | 1.24 | [1.11-1.38] | 1.33 | [1.05-1.8] |
| ^a^ Multivariable conditional logistic regression adjusted for race, education, household income, migration status, metabolic/endocrine conditions status (obesity, diabetes, thyroid disorders and other metabolic disorders), primary care data status, IMD and birth year.  ^b^ Estimation performed only for time intervales with n≥5 of women with both infertility diagnosis and psychiatric conditions. | | | | | | |

**Supplementary Figure 2: Association^a,b^ between female infertility and psychiatric conditions by subgroup and by time point (3-year intervals) relative to the infertility diagnosis.**

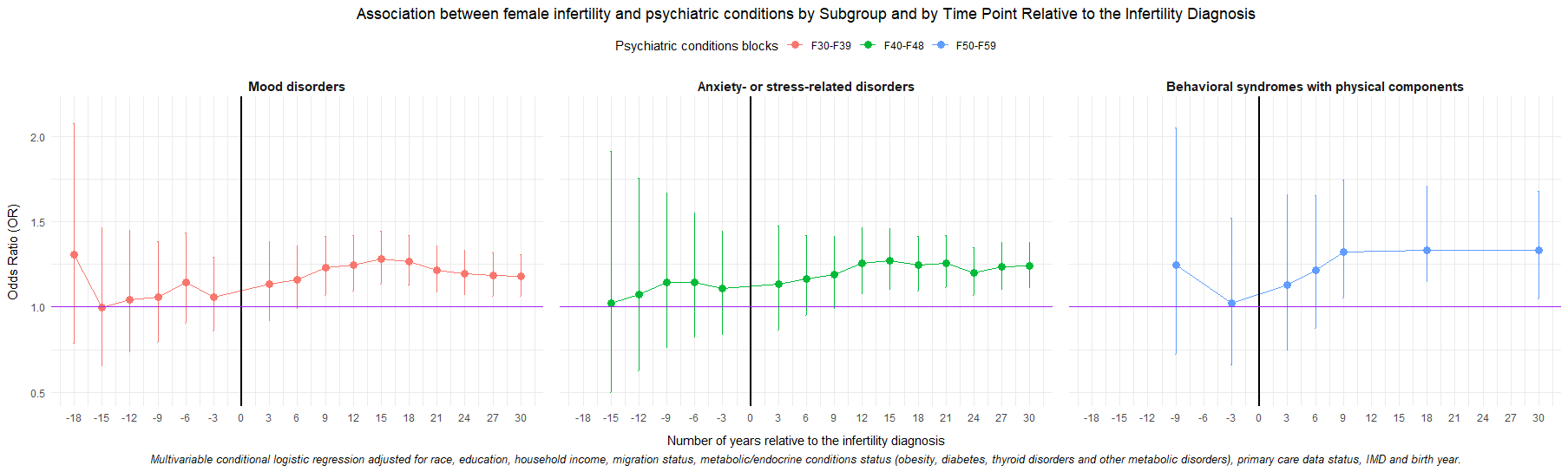
^a^ Multivariable conditional logistic regression adjusted for race, education, household income, migration status, metabolic/endocrine conditions status (obesity, diabetes, thyroid disorders and other metabolic disorders), primary care data status, IMD and birth year.

^b^ Estimation performed only for time intervales with n≥5 of women with both infertility diagnosis and psychiatric conditions.

**Supplementary Table 9: Association^a,b^ between female infertility and psychiatric conditions by time point (1-year intervals) relative to the infertility diagnosis.**

|  | **Mood disorders F30-F39** | | **Anxiety- or stress-related disorders F40-F48** | | **Behavioral syndromes with physical components F50-F59** | |
| --- | --- | --- | --- | --- | --- | --- |
| **Time point** | **OR_adj_** | **95% CI** | **OR_adj_** | **95% CI** | **OR_adj_** | **95% CI** |
| **-3** | 1.06 | [0.86-1.29] | 1.11 | [0.84-1.45] | 1.02 | [0.66-1.52] |
| **-2** | 1.04 | [0.85-1.27] | 1.09 | [0.83-1.41] | Non applicable^b^ | Non applicable^b^ |
| **-1** | 1.04 | [0.86-1.25] | 1.15 | [0.90-1.46] | Non applicable^b^ | Non applicable^b^ |
| **1** | 1.05 | [0.87-1.26] | 1.14 | [0.89-1.44] | 0.99 | [0.67-1.44] |
| **2** | 1.08 | [0.91-1.28] | 1.21 | [0.96-1.50] | 1.07 | [0.75-1.50] |
| **3** | 1.10 | [0.93-1.30] | 1.13 | [0.91-1.39] | Non applicable^b^ | Non applicable^b^ |
| ^a^ Multivariable conditional logistic regression adjusted for race, education, household income, migration status, metabolic/endocrine conditions status (obesity, diabetes, thyroid disorders and other metabolic disorders), primary care data status, IMD and birth year.  ^b^ Estimation performed only for time intervales with n≥5 of women with both infertility diagnosis and psychiatric conditions. | | | | | | |

**Supplementary Figure 3: Association^a,b^ between female infertility diagnosis and psychiatric conditions by time point (1-year intervals) relative to the infertility diagnosis.**

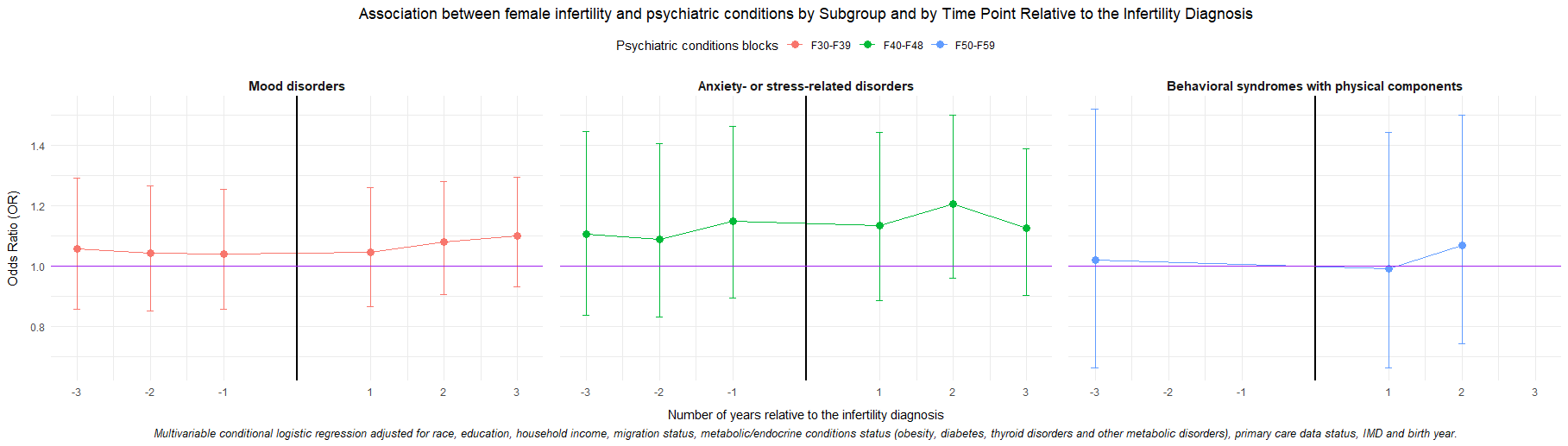

^a^ Multivariable conditional logistic regression adjusted for race, education, household income, migration status, metabolic/endocrine conditions status (obesity, diabetes, thyroid disorders and other metabolic disorders), primary care data status, IMD and birth year.

^b^ Estimation performed only for time intervales with n≥5 of women with both infertility diagnosis and psychiatric conditions.

**Supplementary Table 10: Sensitivity analysis of the association between female infertility diagnosis and psychiatric conditions over the study period and at specific time points (6-year intervals) relative to infertility diagnosis within information source-based subsamples.**

| **Variable** | **All sources except** | **Any time** | **Time point** | | | | | | | |
| --- | --- | --- | --- | --- | --- | --- | --- | --- | --- | --- |
|  |  |  | **-18** | **-12** | **-6** | **6** | **12** | **18** | **24** | **30** |
| **Infertility diagnosis information source** | | | | | | | | | | |
| Mood disorder | Self-report | 1.21[  1.10;1.33] | 1.34[0.81-2.14] | 1.04[0.73-1.45] | 1.15[0.91-1.44] | 1.21[0.96-1.51] | 1.19[1.03-1.37] | 1.3[1.15-1.47] | 1.23[1.1-1.38] | 1.19[1.07-1.33] |
|  | Hospital | 1.22[1.10-1.36] | 1.76[0.93-3.19] | 1.3[0.84-1.95] | 1.31[0.98-1.74] | 1.39[1.05-1.82] | 1.28[1.07-1.53] | 1.38[1.18-1.6] | 1.26[1.1-1.44] | 1.19[1.04-1.35] |
|  | Primary care | 1.22 [1.06-1.41] | 1.1[0.57-2] | 0.88[0.54-1.38] | 1.09[0.79-1.47] | 1.16[0.85-1.56] | 1.19[0.98-1.43] | 1.28[1.08-1.51] | 1.21[1.03-1.42] | 1.18[1.01-1.38] |
| Anxiety- or stress-related disorders | Self-report | 1.20[1.09-1.32] | 0.88[0.33-1.98] | 1.03[0.6-1.7] | 1.15[0.83-1.57] | 1.19[0.88-1.59] | 1.21[1.01-1.44] | 1.26[1.09-1.45] | 1.25[1.11-1.42] | 1.24[1.1-1.39] |
|  | Hospital | 1.28[1.15-1.42] | 1.59[0.5-4.28] | 1.4[0.72-2.55] | 1.24[0.83-1.8] | 1.25[0.86-1.78] | 1.44[1.17-1.76] | 1.47[1.24-1.73] | 1.37[1.18-1.58] | 1.33[1.17-1.51] |
|  | Primary care | 1.12 [0.96-1.30] | 0.45[0.07-1.54] | 1.13[0.55-2.15] | 1.22[0.77-1.86] | 1.3[0.85-1.95] | 1[0.84-1.17] | 1.08[0.87-1.33] | 1.1[0.91-1.33] | 1.09[0.91-1.3] |
| behavioral syndromes with physical components | Self-report | 1.33[1.05-1.67] | 1.83[0.69-4.37] | 1.5[0.74-2.86] | 0.91[0.73-0.82] | 0.95[0.57-1.52] | 1.34[1-1.78] | 1.32[1.01-1.72] | 1.36[1.05-1.74] | 1.33[1.03-1.68] |
|  | Hospital | 1.35[1.05-1.73] | 0.7[0.11-2.69] | 1.28[0.5-2.87] | 0.83[0.44-1.43] | 0.81[0.44-1.41] | 1.42[1.03-1.94] | 1.35[1-1.8] | 1.37[1.03-1.81] | 1.34[1.02-1.75] |
|  | Primary care | 1.38 [0.95-1.97] | 2.97[0.95-8.68] | 1.44[0.51-3.54] | 0.93[0.4-1.93] | 1.05[0.47-2.12] | 1.19[0.75-1.84] | 1.36[0.91-2] | 1.36[0.92-1.98] | 1.31[0.88-1.9] |
| **Psychiatric conditions information source** | | | | | | | | | | |
| Mood disorder | Self-report | 1.25[1.11-1.40] | 1.57[0.94-2.53] | 1.12[0.78-1.56] | 1.19[0.93-1.5] | 1.27[1-1.59] | 1.21[1.04-1.4] | 1.32[1.16-1.49] | 1.23[1.09-1.38] | 1.19[1.06-1.33] |
|  | Hospital | 1.28[1.13-1.46] | 1.3[0.79-2.08] | 1.04[0.73-1.44] | 1.13[0.89-1.42] | 1.21[0.96-1.51] | 1.2[1.04-1.39] | 1.3[1.14-1.47] | 1.25[1.11-1.41] | 1.19[1.06-1.34] |
|  | Primary care | 1.23[1.09-1.40] | 1.29[0.78-2.05] | 1.02[0.71-1.43] | 1.15[0.9-1.45] | 1.23[0.97-1.55] | 1.18[1.01-1.37] | 1.28[1.12-1.46] | 1.19[1.05-1.35] | 1.17[1.03-1.32] |
| Anxiety- or stress-related disorders | Self-report | 1.29[1.16-1.44] | 0.99[0.37-2.26] | 1.13[0.66-1.86] | 1.17[0.85-1.6] | 1.22[0.9-1.63] | 1.2[1-1.44] | 1.28[1.11-1.47] | 1.27[1.12-1.43] | 1.25[1.11-1.4] |
|  | Hospital | 1.21[1.06-1.38] | 0.88[0.33-1.98] | 1.07[0.86-1.75] | 1.11[0.8-1.52] | 1.17[0.86-1.57] | 1.16[0.97-1.39] | 1.26[1.09-1.45] | 1.26[1.11-1.43] | 1.26[1.11-1.41] |
|  | Primary care | 1.18[1.03-1.33] | 0.74[0.25-1.77] | 0.98[0.52-1.72] | 1.01[0.7-1.43] | 1.05[0.74-1.46] | 1.1[0.9-1.34] | 1.18[1-1.39] | 1.21[1.05-1.39] | 1.16[1.01-1.32] |
| behavioral syndromes with physical components | Self-report | 1.37[1.06-1.75] | 1.76[0.67-4.15] | 1.46[0.72-2.77] | 0.89[0.52-1.43] | 0.92[0.55-1.48] | 1.34[1-1.77] | 1.34[1.03-1.74] | 1.38[1.07-1.76] | 1.34[1.05-1.7] |
|  | Hospital | 1.48[1.12-1.95] | 1.62[0.62-3.79] | 1.4[0.69-2.65] | 0.86[0.51-1.39] | 0.91[0.54-1.45] | 1.32[0.99-1.75] | 1.33[1.02-1.72] | 1.37[1.07-1.76] | 1.34[1.05-1.7] |
|  | Primary care | 1.52[1.15-1.99] | 1.16[0.32-3.27] | 1.08[0.46-2.27] | 0.83[0.46-1.42] | 0.91[0.51-1.53] | 1.43[1.04-1.93] | 1.46[1.1-1.93] | 1.44[1.09-1.89] | 1.39[1.06-1.8] |

^a^ Multivariable conditional logistic regression adjusted for race, education, household income, migration status, metabolic/endocrine conditions status (obesity, diabetes, thyroid disorders and other metabolic disorders), primary care data status, IMD and birth year.
